# Application of REDECA Framework to Improve Safety and Health of Agricultural Tractor Drivers

**DOI:** 10.1101/2024.07.10.24310227

**Authors:** Negin Ashrafi, Kamiar Alaei, Greg Placencia, Maryam Pishgar

## Abstract

*Introduction:* Despite tremendous efforts, including research, teaching, and extension, toward improving the safety of agricultural tractor drivers, the number of incidents related to agricultural tractor drivers has not declined. This evidence points out an urgent need to explore artificial intelligence (AI) solutions to improve the safety of tractor drivers. *Methods:* This paper uses 171 Fatality Assessment and Control Evaluation (FACE) reports related to tractor drivers and a new framework called Risk Evolution, Detection, Evaluation, and Control of Accidents (REDECA) to identify existing AI solutions and specific areas where AI solutions are missed and can be developed to reduce incidents and recovery time. Fatality reports of tractor drivers were categorized into six main categories, including run over, pinned by, fall, others (fire and crashes), roll over, and overturn. Each category was then subcategorized based on similarities of incident causes in the reports. *Results:* The application of the REDECA framework revealed potential AI solutions that could improve the safety of tractor drivers. In all categories, the REDECA framework lacks AI solutions for three elements, including the probability of reducing recovery time in R3, detecting changes between R2 and R3, and intervention to send workers to R2. Except for the run over category, all other categories were missing AI solutions for interventions to prevent entry to the R3 element of the REDECA. In addition, the fall, roll over, and overturn categories lacked AI intervention that minimized damage and recovery in R3. *Conclusions:* The outcome of this study shows an urgent need to develop AI solutions to improve tractor driver safety.

## 1. Introduction

Agriculture has the highest rates of fatality incidents in the US [1]. While the number of fatal incidents dropped from around 1000 cases in the early 1990s to less than 600 cases in 2019, this has been because the number of workers has dropped over 30 years. Because of more efficient machinery and systems [2]. Moreover, within all types of agricultural fatal injuries (tractor, roadway, grain bins, farm equipment, Terrain Vehicle (ATV), electrocution, animals, manure storage, and others), the number of tractor-related injuries remains high, with 213 cases from 1999 to 2019 [3]. Over the past few years, several efforts have been made to improve the safety of agricultural workers. These include research, teaching, and extension. In the case of tractor-related injuries, for instance, the focus has been on improving the design and functionality of tractors, while the teaching and extension focus has been on providing farmers safe practices. Despite such efforts, fatal tractor-related incidents are common, indicating a need to explore different approaches to tackle tractor-related injuries.

AI has been applied to many domains. In 2019 alone, over 20,000 papers were published to show the application of AI in various industries [4]. Both academia and industries have used AI to address a variety of issues, including decision-making [5], environmental monitoring [6,7], operational cost reduction [8,9], and productivity [9]. Machine learning algorithms have also been used to detect vocal disorders in workers who frequently use their voices [10], and to detect indoor crews in the event of fire. Gomez-Gil et al. used EMG readings to steer a tractor with almost the same accuracy as manual steering [11]. Szczepaniak et al. developed models to assess the stability and steerability of agricultural machines that could be adapted to drivers’ characteristics to improve safety [12]. Sensors can measure vibrations experienced by farmers using agricultural aircraft. Tri-axial accelerometers were used to measure acceleration at the seat level [13]. Kociolek et al. showed that operators on quad bikes were exposed to head and neck vibration at higher than permissible levels of exposure [14]. Similarly, Calvo et al. used three different accelerometers to measure hand-to-arm vibration and repetitive action (OCRA) levels for farmers who used power tillers. The result indicated vibrational exposure far above acceptable exposure levels [15]. These studies highlight mounting evidence that AI technology can successfully detect, identify, and forecast unsafe behavior in potentially dangerous working environments.

REDECA (Risk Evolution, Detection, Evaluation, and Control of Accidents) [16] is a novel framework developed by the authors to theorize how AI methods can anticipate and control the risk of exposure in a worker’s immediate environment. The REDECA framework is based on the Swiss cheese model [17] that depicts how incidents of injury occur in complex systems. This model conceptualizes multiple layers of defenses and safeguards (or interventions) to prevent incidents. The REDECA framework includes several elements. First, the different states where workers can be. R1 is where workers have minimal to no risk of exposure. R2 indicates exposure to hazard and increased risk of injury. R3 indicates a harmful work-related event. The second is monitoring transitions to adverse states. These can be technologies that predict the probability of transitioning among states. It can also be technologies that detect transitions among states. Finally, there are intervention strategies to keep workers safe or reduce the impact of an adverse event.

The objectives of this paper are: (1) identify root causes of agricultural tractor driver incidents; (2) apply the REDECA framework to determine all steps/ stages involved before and after the occurrence of incidents; (3) determine existing AI solutions to reduce agricultural tractor driver incidents; (4) identifying opportunities for both industry and academia to propose new AI interventions to improve the safety of tractor drivers based on missing REDECA elements; and (5) provide good general practices to improve the safety of tractor drivers.

## 2. Materials and Methods

Agriculture remains the most dangerous occupation in the U.S. (BLS, 2020) [1]. Among all agricultural injuries, tractor-related injuries are the highest [3]. Several databases track all occupational incidents, such as Ag Injury News Clippings, the Fatality Assessment and Control Evaluation (FACE), and the Bureau of Labor (BOL) statistics. We studied FACE reports on fatal incidences among agricultural tractor drivers because they were the most complete and comprehensive.

We used the following procedures to extract and analyze FACE report data. (1) We accessed FACE reports at national and state levels from the Center for Disease Control and Prevention (CDC) website, (2) using the keywords “agriculture” to find agricultural-related reports, (3) and “machine farming” to find machine-related cases. (4) We extracted and saved the reports to an Excel file. (5) We then identified all tractor-related cases by searching the Excel file for the keyword “tractor”. (6) In cases where national and state reports were identical, we analyzed the cases as a single incident. All reports were reviewed, and unwitnessed reports were excluded from analysis.

Further analysis (7) categorized reports based on types of tractor-related incidents: roll over, run over, overturn, pinned by, fall, and others that include fire, crashed incidents, as well as other types of incidents. Additionally, (8) in each category, reports with the same causes were sub-categorized together. Categories and subcategories are detailed in Table 1.

**Table 1.** Incident categories and sub-categories.

| Types of Categories | Types of Sub- Categories |
| --- | --- |
| Run Over | (1) Run over due to unsecured seat placement. |
|  | (2) Run over due to the tractor left in gear after its last use. |
|  | (3) Run over because of the weight of the water tank trailer and high speed. |
|  | (4) Run over because the tractor engine was operating at high rpm, slipped into gear, and accelerated forward at high speed. |
|  | (5) Run over due to the wheels on the right side of the tractor running over a tree stump. |
|  | (6) Run over due to the tractor being in forward gear and the driver standing on the ground trying to turn the tractor on. |
|  | (7) Run over due to tractor being left on a hill while running without driver and dismounted from its initial attached loader bucket. |
|  | (8) Run over due to driver having a seizure and falling out of the tractor because they did not use a seat belt. |
|  | (9) Run over due to unintended door opening causing the driver to fall out of the tractor. |
|  | (10) Run over due to non-functional brake. |
|  | (11) Run over due to non-functional gear. |
|  | (12) Run over due to the driver falling off from the tractor moving at high speed. |
|  | (13) Run over due to a driver falling off the tractor while moving. The driver tried to jump on the tractor, slipped and engaged the gear making the tractor move while the victim was on the ground. |
|  | (14) Run over due to falling off the tractor while making a sharp turn. |
|  | (15) Run over due to poor driver visibility. |
| Pinned by | (1) Pinned between the enclosed auger and the tractor steering wheel due to an accident between the tractor and barn wall. |
|  | (2) Pinned to tree while inside tractor cab. |
|  | (3) Pinned by a truck bed attached to a tree by a chain. |
|  | (4) Pinned by and compressed by an attachment while trying to detach it. |
|  | (5) Pinned by a tractor that drove into a ditch where the victim was located. |
|  | (6) Pinned by hayrack roll over that pinned the victim to the left rear tractor tire. |
| Fall | (1) Fall out of the tractor while driving due to an unattached seat. |
|  | (2) Fall off due to slip on a tractor step. |
|  | (3) Fall off due to tractor driving at high speed on a incline and driver not wearing a seatbelt.<br>(4) Fall off from tractor due to heart issues and not wearing a seatbelt.<br>(5) Fall off due to tractor jumping jack-knifed causing the driver to be thrown off from the tractor.<br>(6) Fall off from tractor due to loss of control while driving. |
| Other category including fires and crashes | (1) Victim struck by hay bale that fell out of elevated bucket.<br>(2) Victim fell while re-attaching a shaft to a tractor due to control lost in the hollow.<br>(3) Entrapment due to victim's shirt entrapped inside an auger not equipped with guard.<br>(4) Entanglement due tractor left on and hay baler left on downward slope.<br>(5) Incident due to a tractor driver hauling a tree higher than the recommended height.<br>(6) Crash due to a semi-truck hitting a tractor causing both victims to be ejected from their respective vehicles.<br>(7) Crash due to a detached wagon moving forward, crushing the user against the tractor.<br>(8) Fire due to the victim puncturing an above ground gas line with a tractor due to poor visibility.<br>(9) Fire due to vinyl shrouds bursting into flames, spreading to the victim's clothing.<br>(10) Fire due to ignition of a tractor struck by a tree. |
| Roll Over | 1) Roll over due to a tractor's sharp turn into a ditch.<br>2) Roll over due to tractor driving and reaching a steep roadside.<br>3) Roll over due de-attaching a wagon from the tractor on a hilly, sharply curved road.<br>4) Roll over due to an attached overweight hay bale and driving on a slope.<br>5) Roll over due to tractor driving on an irregular surface causing its center of gravity to shift resulting in tractor instability.<br>6) Roll over due to a rasied tractor bucket and attempting to make a left turn while on the incline.<br>7) Roll over due to driver foot slipping off the tractor's clutch while trying to pull out another tractor stuck in mud.<br>8) Roll over due to the tractor being stopped and dismounted without setting the brakes and leaving the manual transmission in gear.<br>9) Roll over due to the tractor not equipped with ROPS and seat belt reaching an embankment while the brakes were not engaged. |
| Overturn | 1) Overturn due to wet/snow/muddy hill causing the tractor to slide down.<br>2) Overturn due to tractor driving on slope terrain and ditch.<br>3) Overturn due to tractor driving on the right side of the road with a heavy trailer was connected and attempting to overcorrect.<br>4) Overturn due to the tractor turning on a slope/hill.<br>5) Overturn due to the tractor sliding down a steep embankment.<br>6) Overturn due to the tractor attached to overweight objects.<br>7) Overturn due to the tractor wheel coming off the ground while driving.<br>8) Overturn due non-functional gear shift.<br>9) Overturn due tractor trying to pull out another tractor stuck into mud.<br>10) Overturn due to speeding tractor. |

We identified each state of work using the REDECA framework – from when tractor drivers started working until the incident happened (R1, R2, R3). We then indicated AI technologies that could predict the probability of transitioning among states, detect transitions among states, and indicate intervention strategies to keep the tractor drivers safe or reduce recovery times when incidents occur. Finally, we found existing AI technologies used by other industries and advised them to be used in the agriculture industry to improve the safety and health of tractor drivers. Figure 1 outlines our methodology.

**Figure 1.**
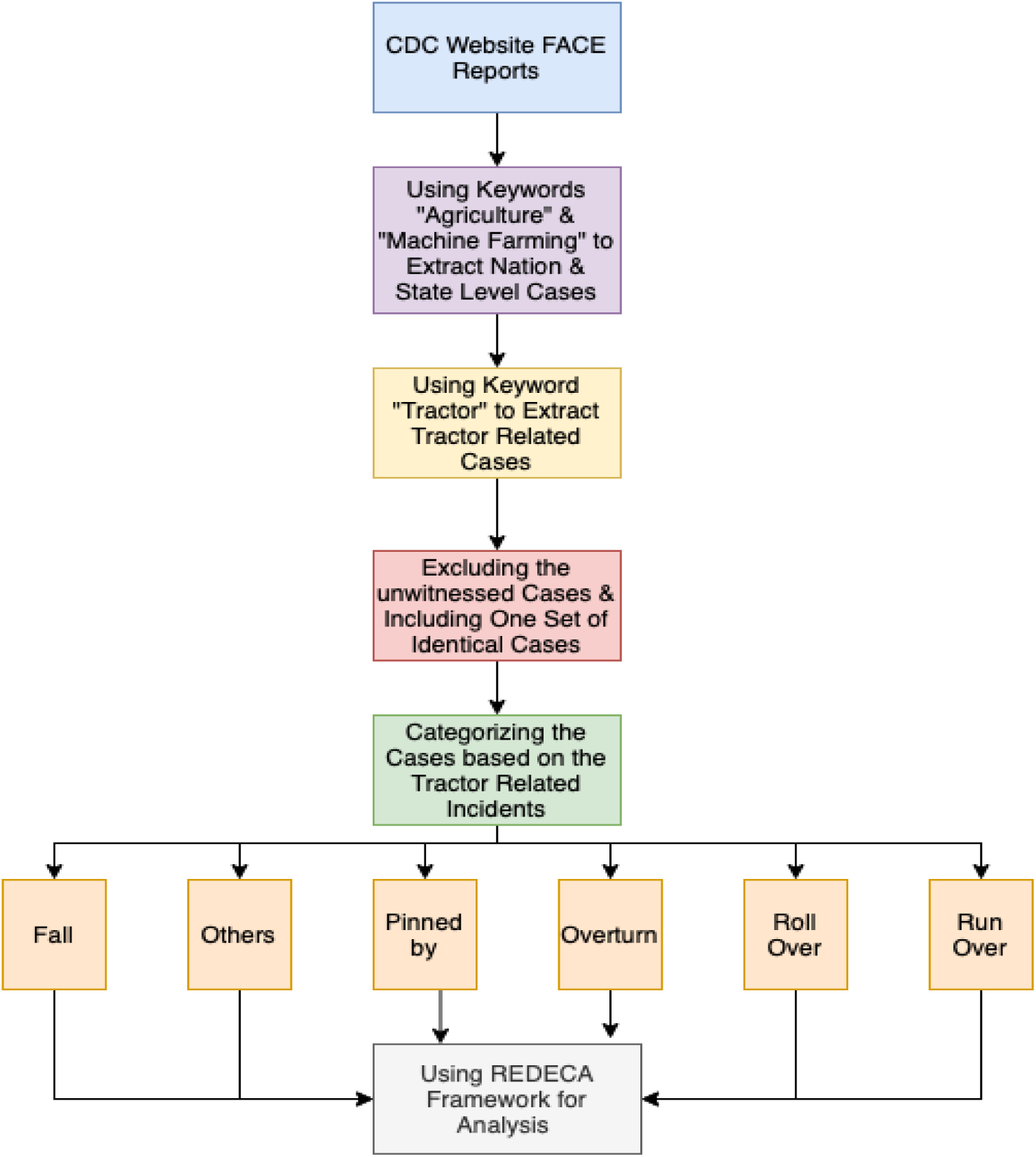
Overall view of the methodology.

## 3. Results

From 442 initial cases of fatalities from FACE reports at national and state levels, we identified 188 tractor-related incidents using the keyword “tractor.” 9 cases were unwitnessed, and 16 were duplicates we combined into single cases. This left 171 cases for analysis.

We subcategorized 160 cases by the kind of tractor-related occurrence. 52 cases were run over, 44 cases of roll over, 7 of pinned by, 42 cases of overturned, 10 cases of fall, and 16 cases of others, including fires, and crashes. We further subcategorized incidents with similar causes: run over had 15 sub-categories, pinned by and fall both had 6, others and overturn both had 10 and roll over had 9.

We analyzed all cases using the REDECA framework (see Appendix A, Tables A1–A7). For example, from reviewing 52 cases of run over, we obtained 15 subcategories, each with different occurrences. We found only one report of subcategories 1, 3, 5, 8, 9, 13, and 15; and two reports of subcategories 2, 4, 7, 11, and 14 respectively. There were also 3 reports for subcategory 10 and 4 reports for subcategory 12, respectively. Finally, there were 21 reports of subcategory 6.

Based on Table A1 (Appendix A), 15 AI solutions exist to measure the probability of entering R3, 2 AI solutions to detect change between R2 to R3, 11 AI interventions to prevent entry to R3, and 4 AI interventions to minimize damage and recovery in R3. However, AI technology cannot establish the probability of reducing recovery time in R3 and detecting transitions between R2 and R3, or intervention to send workers to R2.

In the case of pinned, we created 6 subcategories from the 6 cases we reviewed. Each subcategory was reported 1 time respectively. Based on Table A2 (Appendix A), 6 AI solutions exist to measure the probability of entering R3. However, there are no AI solutions for the probability of reducing recovery time for R3, detecting change between R2 to R3, intervention to prevent entry to R3, Intervention to send workers to R2, and intervention to minimize damage and recovery in R3. There are 6 AI interventions to prevent entry to R3.

In case of fall, we created 6 subcategories from the 6 cases reviewed, with 1 report per case except one classification with 2 reports. Based on Table A3 (Appendix A), there are 6 existing AI solutions to measure the probability of entering R3. However, there are no AI solutions for the probability of reducing recovery time for R3, detecting the change between stages R2 to R3, intervention to send workers to R2, and intervention to prevent entry to R3. There are 2 AI interventions to prevent entry to R3. Finally, there is only 1 AI intervention to minimize damage and recover.

In cases of “others,” we created 10 subcategories from the 10 cases we reviewed, each with 1 reported case. Based on Tables A.4 and A.6, AI solutions exist to measure the probability of entering R3. However, there are no AI solutions for the probability of reducing recovery time in R3, detecting change between R2 to R3, nor intervention to prevent entry to R3, nor to send workers to R2. There are 7 AI interventions to prevent entry to R3. Finally, there are no AI interventions to send workers to R1 and to minimize damage, but there are 2 AI interventions to minimize damage and recovery.

In cases of roll over, we obtained 9 subcategories from the 44 cases we reviewed. Each classification had different numbers of reports: classes 1, 3, and 5 contain 6 reports, respectively. Class 4, 6, and 9 contain 3 reports respectively. Finally, classes 2, 7, and 8 include 4, 1, and 2 reports, respectively.

Based on Table A5 (Appendix A), only 5 AI solutions exist to measure the probability of entering R3 and 5 AI interventions to prevent entry to R3. Except where the AI solutions are not applicable within the context of a tractor driver, there are no existing AI solutions applicable to all scenarios of the REDECA framework. There are no AI solutions for the probability of reducing recovery time in R3, detecting change between R2 to R3, intervention to prevent entry to R3, intervention to send workers to R2, and intervention to minimize damage and recovery in R3.

In cases of overturn, we created 10 classifications from the 42 cases we reviewed. Each classification had different numbers of reports: classes 3, 7, 8, 9, and 10, each reported 1 time, respectively. Class 1, 2, 4, 5, and 6 had 10, 12, 4, 11, and 6 reports respectively. Based on Table A6 (Appendix A), only 5 AI solutions exist to measure the probability of entering R3 and 5 AI interventions to prevent entry to R1. Except where an AI solution is not applicable within the context of a tractor driver, there are no existing AI solutions applicable to all scenarios of the REDECA framework. There are no AI solutions for the probability of reducing recovery time in R3, detecting change between R2 to R3, intervening to prevent entry to R3, sending workers to R2, or minimizing damage and recovery in R3.

## 4. Discussion

The application of AI to several industrial domains has been described as the Fourth Industrial Revolution [18]. Innovations in artificial intelligence using sensors, robots, and ML algorithms have been shown to increase productivity and improve the safety and health of workers in the workplace. In agriculture, despite tremendous efforts to improve safety, the number of tractor-related incidents remains high [3]. REDECA is a state-of-the-art framework that identifies work states from the start of the workflow until an incident occurs (R1, R2, R3). It then categorizes AI use in OSH to highlight strengths, opportunities, and weaknesses clearly and efficiently.

In this paper, we extracted tractor–related fatality incidents from FACE reports on the CDC website. We then used the REDECA framework to assess processes before incidents and identified potential AI solutions that can reduce these incidents.

For run over cases, class 6, that is, “driver run over by a tractor in gear, while the driver was standing on the ground starting the tractor, and delay in receiving care,” had the most reports. (21). The frequency of such cases indicates a critical need to develop AI solutions to reduce their occurrence. Moreover, our REDECA framework analysis of run over cases, highlighted a lack of AI technology to reduce the probability of recovery time in R3, to detect changes between R2 and R3, and interventions to send workers to R2. As a result, researchers and industry have an opportunity to address said issues.

After analyzing FACE reports, we developed the following recommendations to reduce run over incidents and consequently improve the health and safety of tractor drivers. 1) Install sensors to alert drivers to turn off tractors before leaving the tractor. 2) Install sensors to alert drivers not to turn on the tractor while they are on the ground. 3) Utilize smart technology to keep tractors in a parked state while in a maintenance state. 4) Install pressure sensors to inform drivers not to leave their seats while the tractor is running. 5) Install sensors to prevent tractors from starting while not in neutral. 6) Equip tractors to make sure seat belts are engaged while the tractor is in operation and install roll over protective structures. 7) Tractor transmissions should always be put in the park before drivers’ dismount to hook up or adjust equipment.

In brief, for pinned cases, there is a lack of AI technology to detect change between R2 to R3, minimize damage and recovery in R3, reduce the probability of recovery time for R3, prevent entry to R3, and send workers to R2. Hence, researchers and industries can focus on developing missing AI technology that improves the safety of tractor drivers.

In brief, for fall cases, there is a lack of AI technology to detect change between R2 to R3, minimize damage and recovery in R3, reduce the probability of recovery time for R3, prevent entry to R3, and send workers to R2. Thus, there is a need for academic and industrial research to develop such AI technologies.

To reduce fall incidents, we suggest: 1) Owners/operators of tractors should ensure that the tractor seat is in good condition and firmly attached to the base. (Vibration sensors or any sensors could alert the user when seats are not properly attached to the base or raised) 2) Tractor manufacturers should give more attention to the safe design of steps and handrails to further increase operator safety. 3) Tractor operators should pay close attention to symptoms of illness and should seek prompt medical attention.

For other cases including fire and crashes, there is a lack of AI technology to detect change between R2 to R3, to reduce the probability of recovery time for R3, to prevent entry to R3, and to send workers to R2. Therefore, developing new AI technology is crucial for these missing elements of the REDECA framework.

To reduce pinned by incidents, we suggest the following: 1) Ensure adequate rest and minimize distractions while driving. 2) Use less busy alternate routes when available when operating agricultural equipment on the road, especially during high traffic times. 3) Install side view mirrors and construct/purchase appropriate warning lights for when the tractor operates on the road. These can be temporarily attached to the tractor as needed. 4) MIFACE recommends that the tractor PTO stub shaft be always guarded to prevent entanglement. 5) operators should not wear loose-fitting clothing when operating farm machinery. Clothing manufacturers should, therefore, consider developing work clothes that tear away in case of entanglement and label clothing when tear resistant. 6) Make smooth clutch or brake pedals non-slip to improve foot pedal control. 7) Drivers should carry a reliable 2-way communication device for emergency communication in case of injury and emergency situations.

For roll over cases, class 1 is “roll over due to tractor sharp turning into a ditch. No ROPS and seat belt”, class 3 is “roll over” due to de-attaching the wagon from the tractor in a hilly road and sharp curve road. No ROPS and seat belt”, and class 5 is “roll over due to tractor driving on an irregular surface causing its center of gravity to shift which resulting in tractor instability.” Class 5 is the most common, with six reports. This implies such cases are critical, and AI solutions should be developed to reduce their occurrence.

Based on the REDECA framework analysis for roll over cases, there is a lack of AI technology for the probability of reducing recovery time in R3, detecting changes between R2 and R3, and intervention to send workers to R2, intervention to prevent entry to R3, and intervention to minimize damage and recovery in R3. New AI technologies are welcome to address these needs.

In brief, for overturn cases, class 1 is “overturn due to wet/snow/muddy hill causing the tractor to slide down. No ROPS or seatbelt”, class 2 “overturn due to tractor driving on slope terrain and ditch. No ROPS or seatbelt”, and class 5 “overturn due to tractor sliding down a steep embankment.” These contain 10, 12, and 11 reports respectively. Given the high negative impact of these cases, it is urgent to develop AI solutions.

Moreover, based on the REDECA framework analysis for roll over cases, there is a lack of AI technology for the probability of reducing recovery time in R3, detecting changes between R2 and R3, and intervention to send workers to R2, intervention to prevent entry to R3, and intervention to minimize damage and recovery in R3. New AI technologies are needed to fulfill the missing REDECA framework elements.

To reduce roll over incidents we recommend the following: 1) Tractor operators should be trained to recognize and understand the hazards associated with towing items that exceed the weight of the tractor. 2) When driving a tractor on a public road, the driver should maintain a safe and well-defined position on the road in the correct traffic lane. The tractor-operator should not pull off the road to allow traffic to pass unless there is a safe and stable location to maneuver. 3) Provide personal communication devices to workers assigned to remote worksites. 4) front-end counterweights should be used to improve traction and stability. 5) Foreign farm laborers should be trained in their native language to operate farm machinery safely and to warn them about local terrain hazards. 6) Tractor front-end loader operators should be warned of overturn hazards and methods to reduce these hazards, including safe driving on sloped ground, changes to the center of gravity caused by a loader bucket, keeping the bucket low while driving, and using counterweights on the tractor. 7) Warn tractor operators about the dangers of fatigue and weariness and have them take frequent breaks.

We observed several common causes across all these types of incidents, for which we recommend the following: 1) Tractor drivers should always use seat belts, and tractors should be equipped with ROPS. 2) Have a trained mechanic inspect used equipment prior to use to ensure equipment has all safety features intact and to note any equipment modifications that may affect equipment performance and function. 3) Routinely inspect tractors to identify potential safety issues, such as old/faded SMV emblems, missing PTO master shields, and roll over protective structure (ROPS) availability. Install/re-install missing or damaged items. 4) Ensure medical conditions are managed by all workers on the farm. 5) Tractor operators should always maintain safe operating speeds. 6) Survey work sites to identify hazards and warn all employees about these hazards. Employees should be encouraged to report any unsafe work conditions as well. 7) Assign age-appropriate tasks to working youth. 8) Farmers, rural residents, and county/state road departments should pursue grading changes or post warning signs along the roadway to alert drivers of dangerous intersections with farm lanes or driveways. 9) Operators should lock both brake pedals together before driving in slippery conditions.

While we have demonstrated the utility of REDECA in identifying hazards when operating tractors, we acknowledge several limitations of the study. For one, developing the probabilities for transitions among states R1, R2, and R3 can be time-intensive and difficult. Out of 442 reported cases, we could only use 171 that were witnessed and not duplicated. It is unclear how many cases go unreported that could skew observations. In addition, the cost of developing AI solutions could be both cost-prohibitive and technologically infeasible. Moreover, it is unclear how well such solutions would integrate into current work processes and whether they would be perceived as disruptive despite potential benefits to safety and occupational health.

For example, one of the authors of this study was personally involved in a research study observing the initial sociotechnological integration of positive train control (PTC) immediately after the Rail Safety Improvement Act of 2008 was enacted. [36] Despite the benefits of using PTC to detect and prevent several severe railway hazards using AI, there was massive pushback against implementing it. For one, the cost to implement was approximately the same as the rail maintenance budget during the time of implementation. The technology also required many years to develop and verify, including establishing the infrastructure to operate the system. Rail culture also needed to change from very punitive to collegial before train operators trusted the system. Despite these, PTC was integrated into passenger rail and continues to operate as planned. The system also gathers data that can be used to establish hazards.

Given the potential benefits of using AI in tractor operations, we encourage future researchers to examine the hazards we have identified. These include how to integrate technologies that can gather data about hazards in tractor operations cost-effectively, with minimal process disruptions. The potential savings preventing injury would be well worth the endeavors.

## 5. Conclusions

AI technology has been successfully spread across all industries, including agriculture, mining, transportation, construction, and more. The use of AI technology has shown significant improvement in the health and safety of the workers in the workplace. REDECA is a useful framework that highlights AI/OSH strengths and opportunities for advances in sensors, robotics, and machine learning algorithms to improve working conditions in the workplace. Given the high and steady fatal incident rate within agricultural workplaces related to tractor drivers, we used the REDECA framework to deeply understand every step involved before and after the occurrence of the incidents among tractor drivers and to identify existing AI solutions used to improve the safety of the tractor drivers. Moreover, the REDECA framework helped identify specific areas where AI solutions are missed and can be developed to reduce the occurrence of incidents and recovery time. There is an urgent need to apply AI solutions to improve the health and safety of agricultural tractor drivers.

## Data Availability

All relevant data underlying the findings of this study are available. Additionally, the data from the Fatality Assessment and Control Evaluation (FACE) reports used in this study are publicly available from the National Institute for Occupational Safety and Health (NIOSH) FACE program website.

## Author Contributions

N.A., M.P.: Involved in all aspects of this study. G.P.: Revision of the manuscript. Expert insights were provided by K.A. All authors have read and agreed to the published version of the manuscript.

## Institutional Review Board Statement

This project did not involve any human subject research.

## Acknowledgments

Its contents are solely the responsibility of the authors and do not necessarily represent the official views of the Centers for Disease Control and Prevention or the Department of Health and Human Services.

## Appendix A.

**Table A.1.**
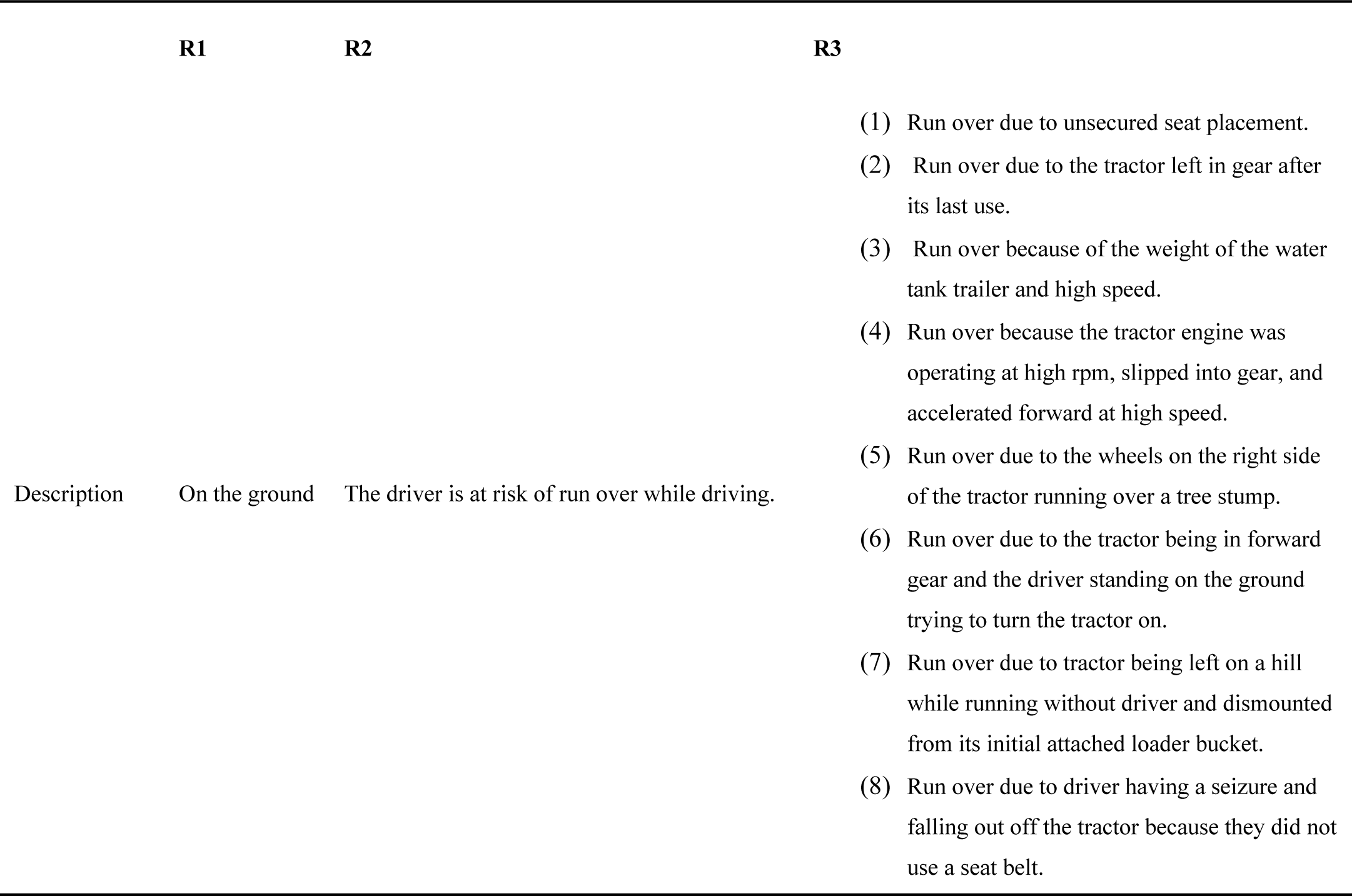
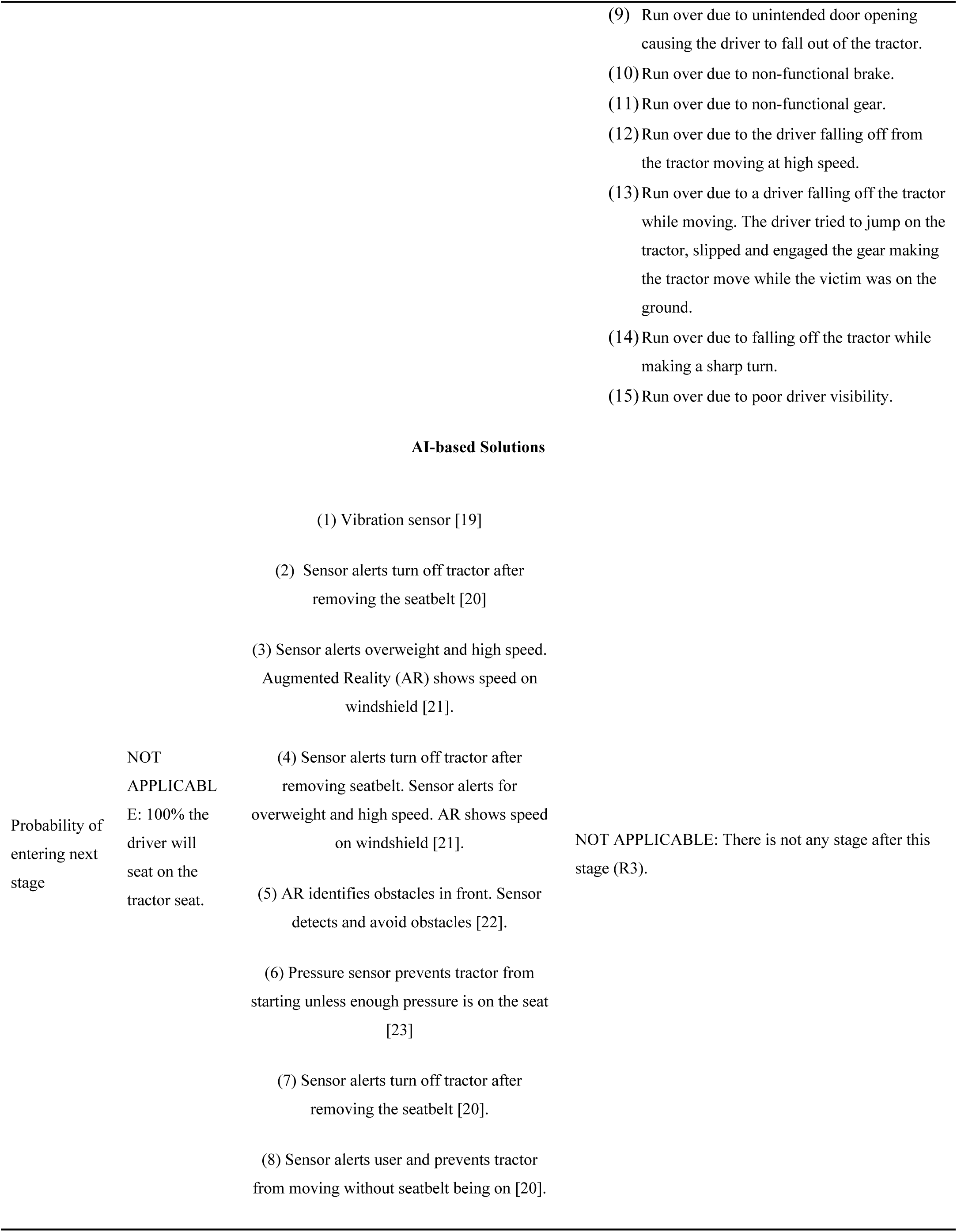
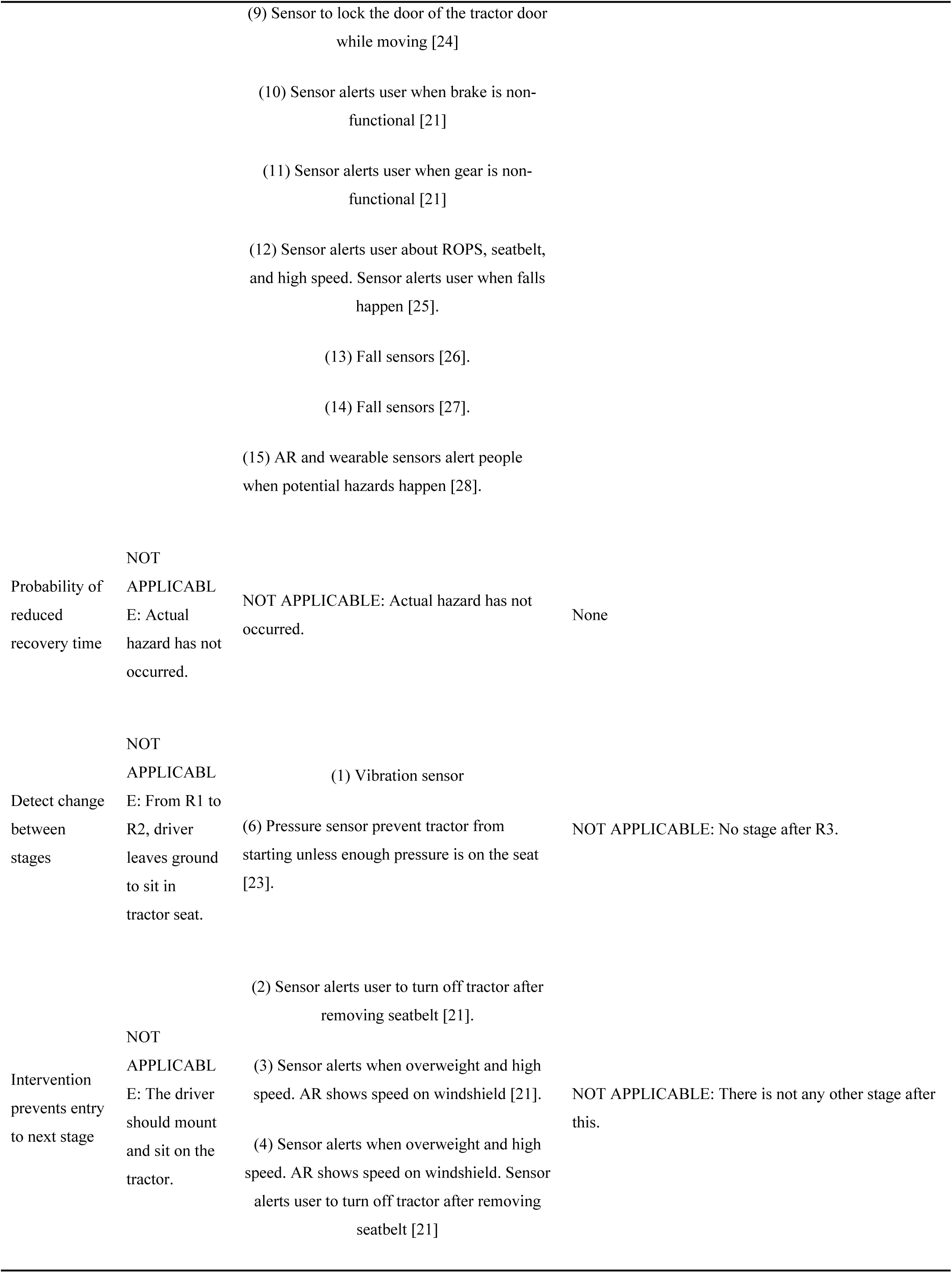
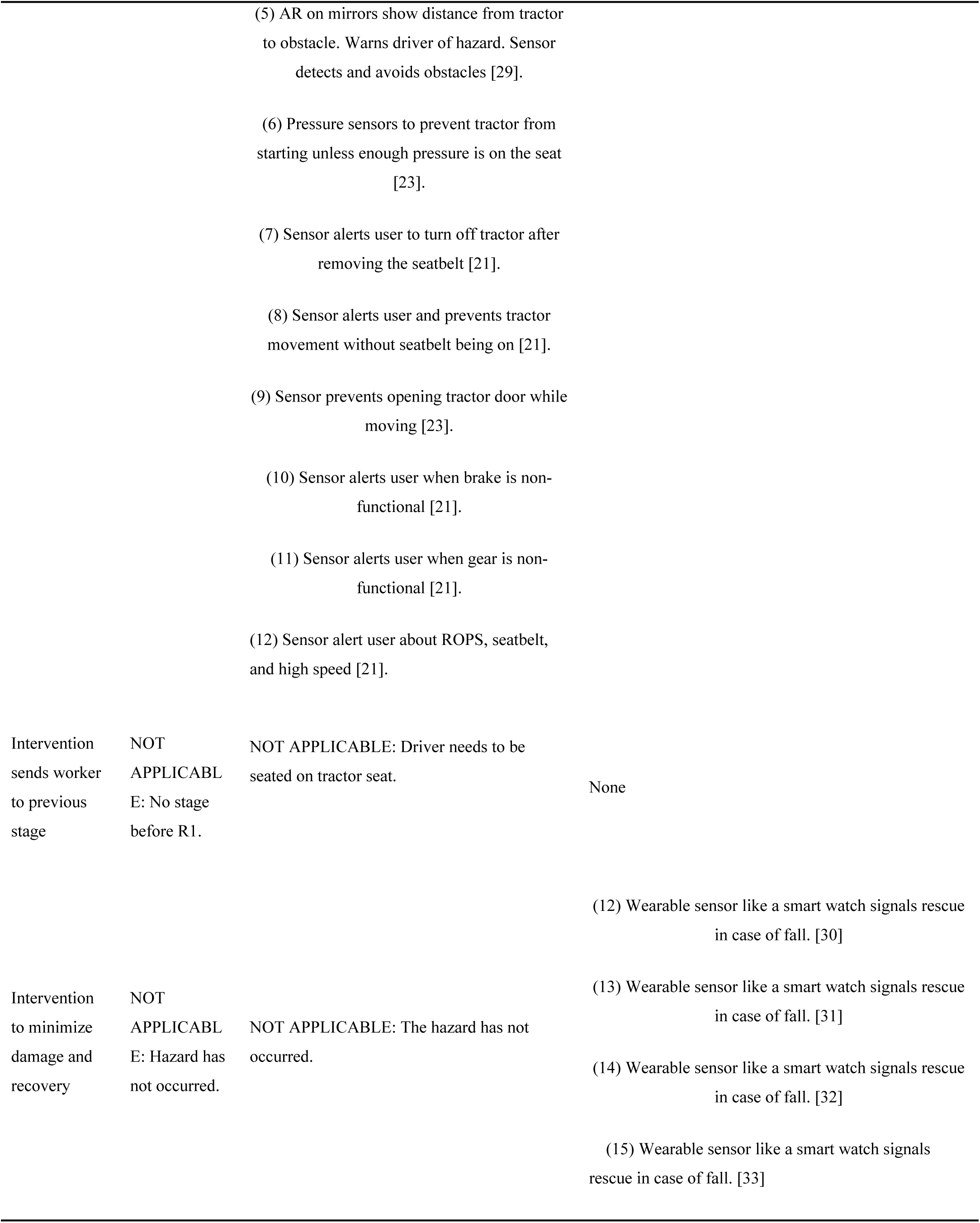
REDECA framework results summary for run over cases.

**Table A.2.** REDECA framework results summary for Pinned by cases.

|  | **R1** | **R2** | **R3** |
| --- | --- | --- | --- |
| Description | On the ground | Driver at risk of being pinned while driving. | (1) Pinned between the enclosed auger and the tractor steering wheel due to an accident between the tractor and barn wall.<br>(2) Pinned to tree while inside tractor cab.<br>(3) Pinned by a truck bed attached to a tree by a chain.<br>(4) Pinned by and compressed by an attachment while trying to detach it.<br>(5) Pinned by a tractor that drove into a ditch where the victim was located.<br>(6) Pinned by hayrack roll over that pinned the victim to the left rear tractor tire. |
| **AI-based Solutions** |  |  |  |
| Probability of entering next stage | NOT APPLICABLE: 100% driver sits on tractor seat. | (1) AR and perception sensors [51]<br>(2) AR and perception sensors [52]<br>(3) AR and perception sensors [55]<br>(4) AR and perception sensors [62]<br>(5) AR and perception sensors [51]<br>(6) AR and perception sensors [52] | NOT APPLICABLE: No stage after R3. |
| Probability of reduced recovery time | NOT APPLICABLE: Hazard has not occurred. | NOT APPLICABLE: Hazard has not occurred. | None |
| Detect change between stages | NOT APPLICABLE:<br>From R1 to R2, the driver leaves ground to sit on tractor seat. | None | NOT APPLICABLE: No stage after R3. |
| Intervention to prevent entry to next stage | NOT APPLICABLE:<br>Driver mounts tractor and sits in it. | None | NOT APPLICABLE: No stage after this. |
| Intervention to send worker to previous stage | NOT APPLICABLE:<br>No stage before R1. | NOT APPLICABLE:<br>Driver needs to sit in tractor seat. | None |
| Intervention to minimize damage and recovery | NOT APPLICABLE:<br>Hazard has not occurred. | NOT APPLICABLE:<br>Hazard has not occurred. | None |

**Table A.3.**
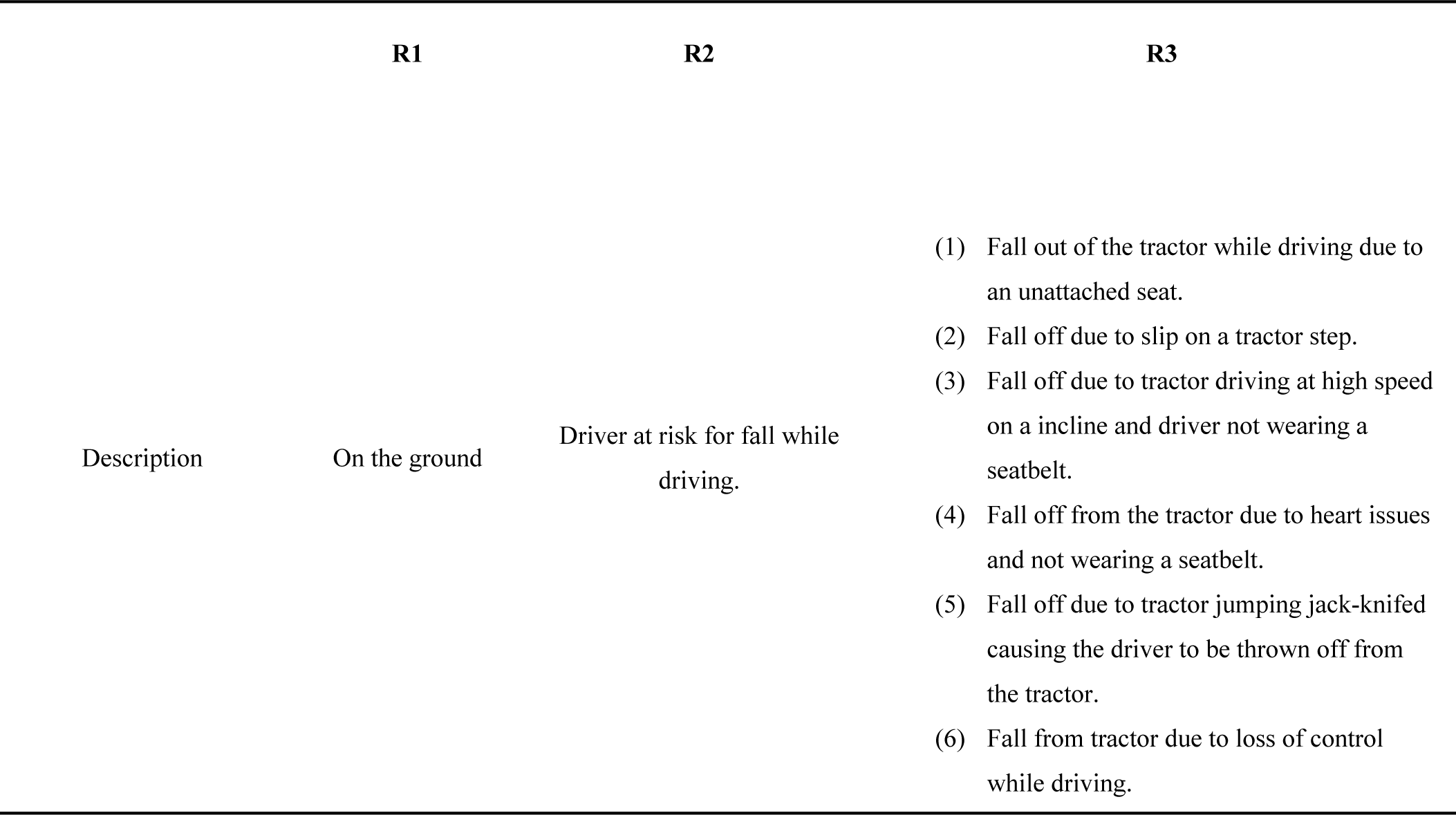
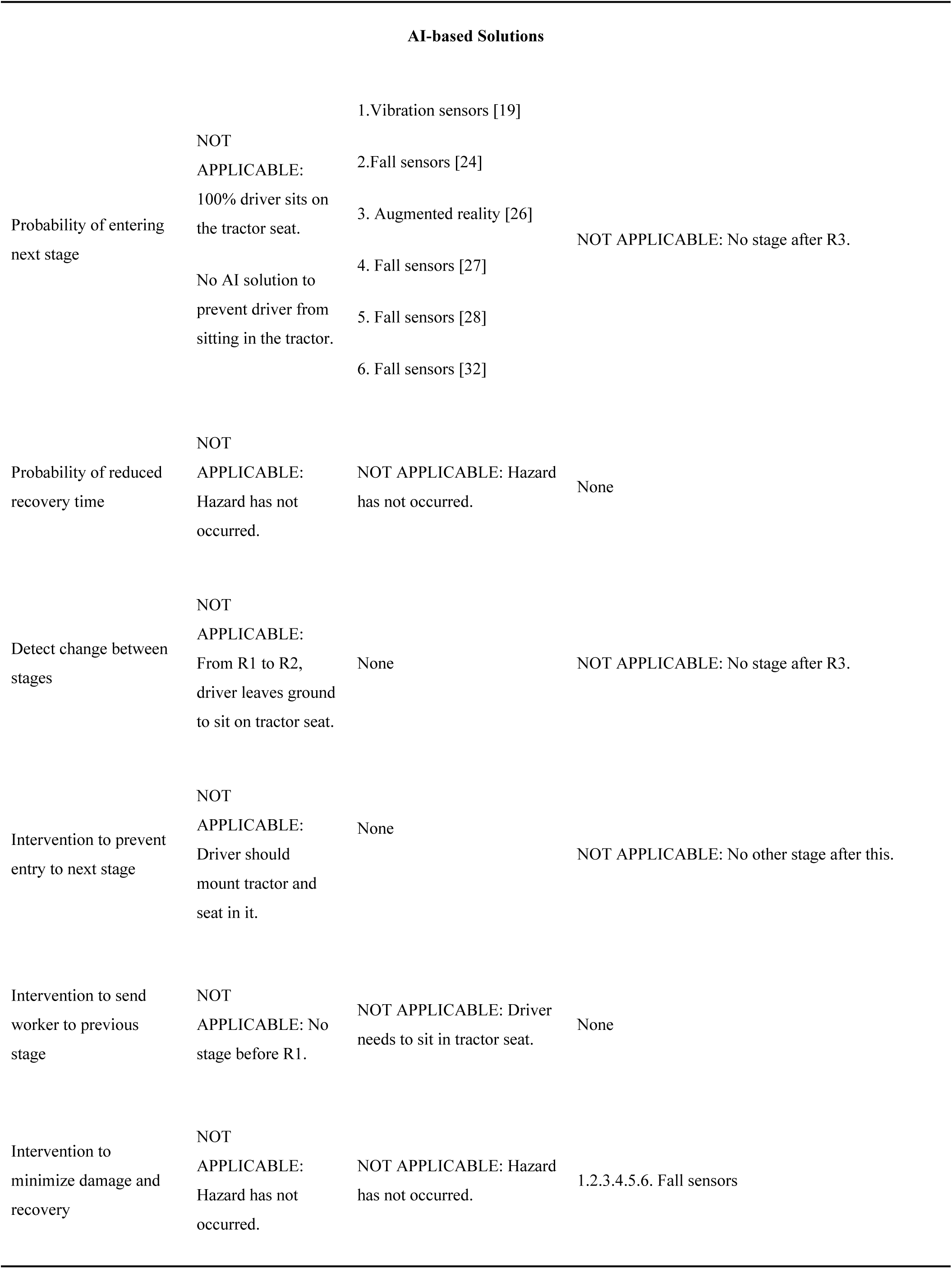
REDECA framework results summary for fall cases.

**Table A.4.**
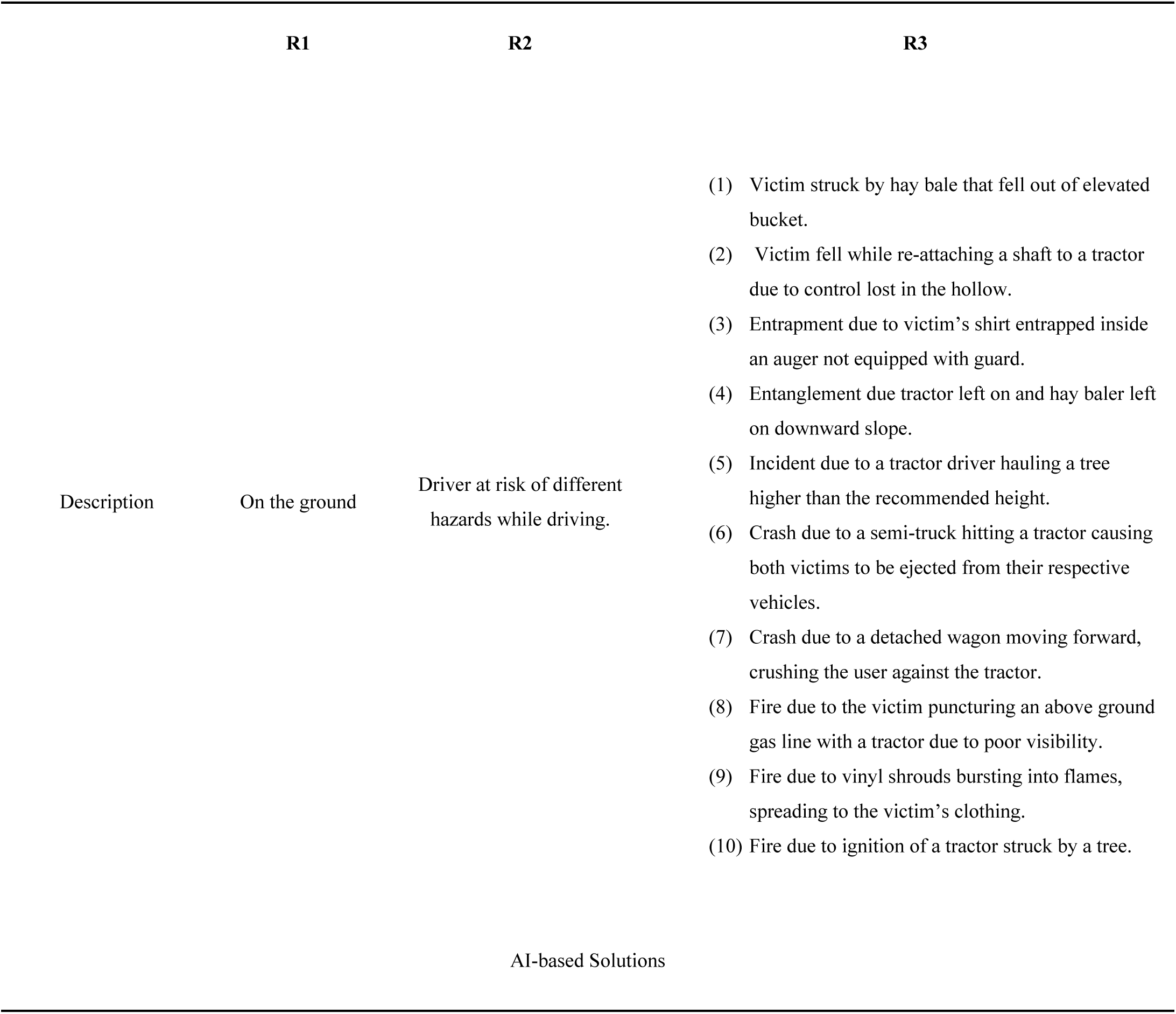
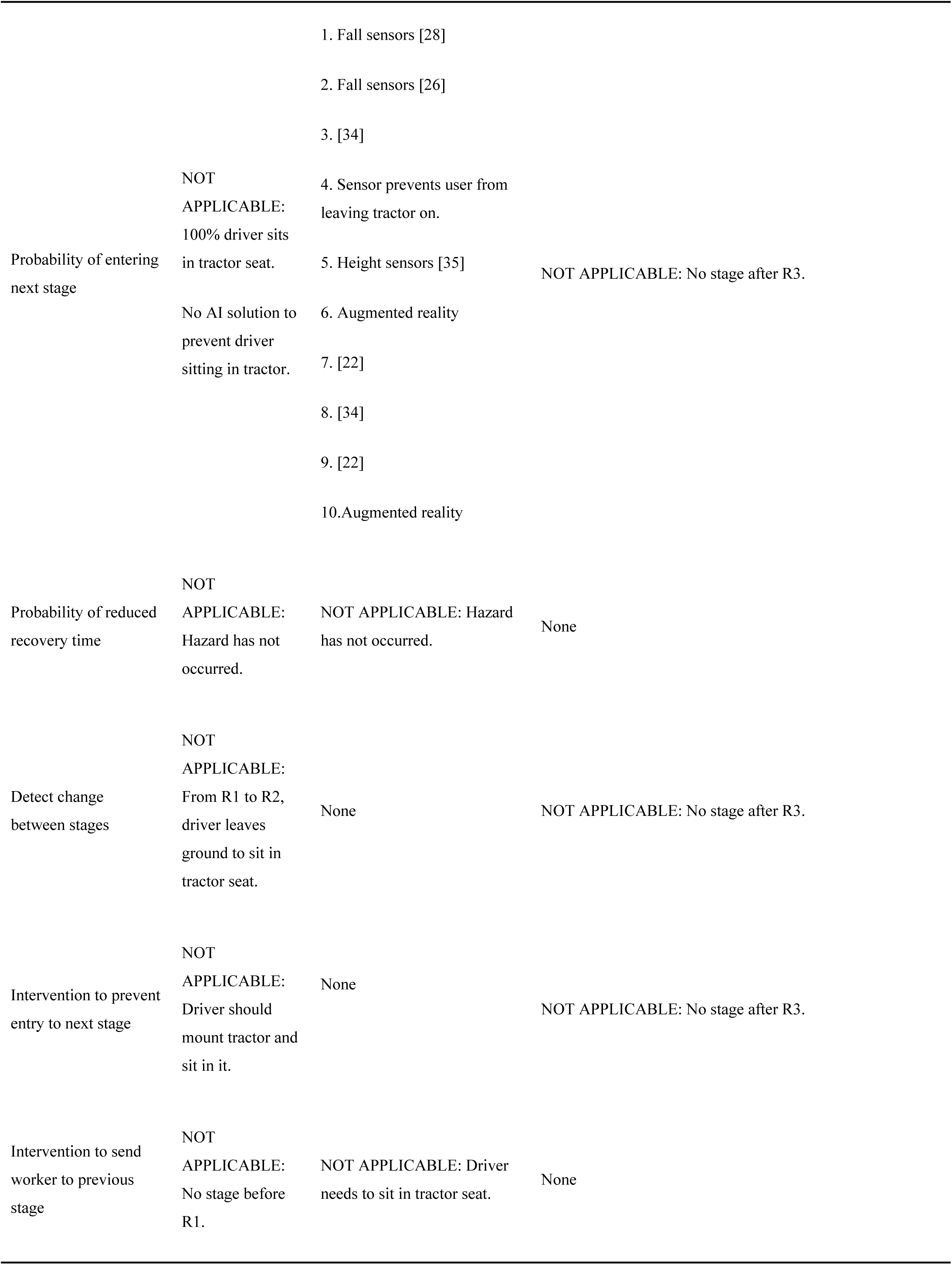
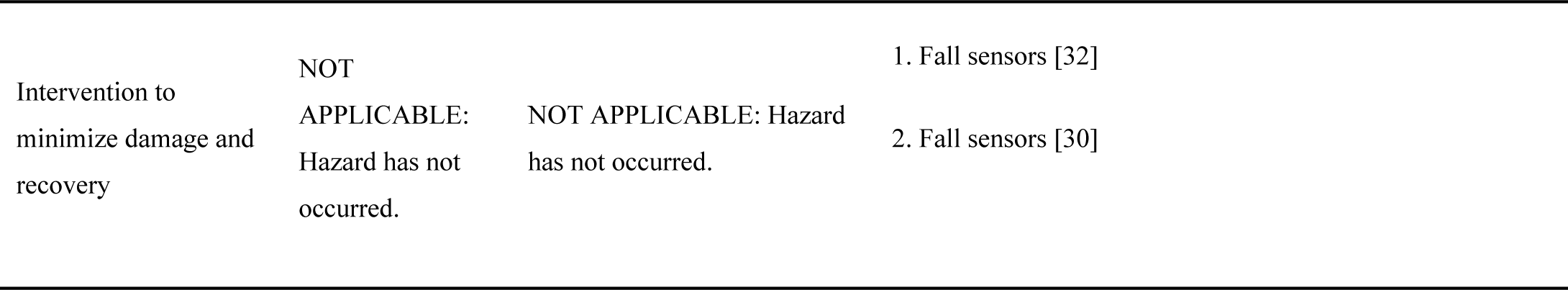
REDECA framework results summary for others including fires and crashes.

**Table A.5.**
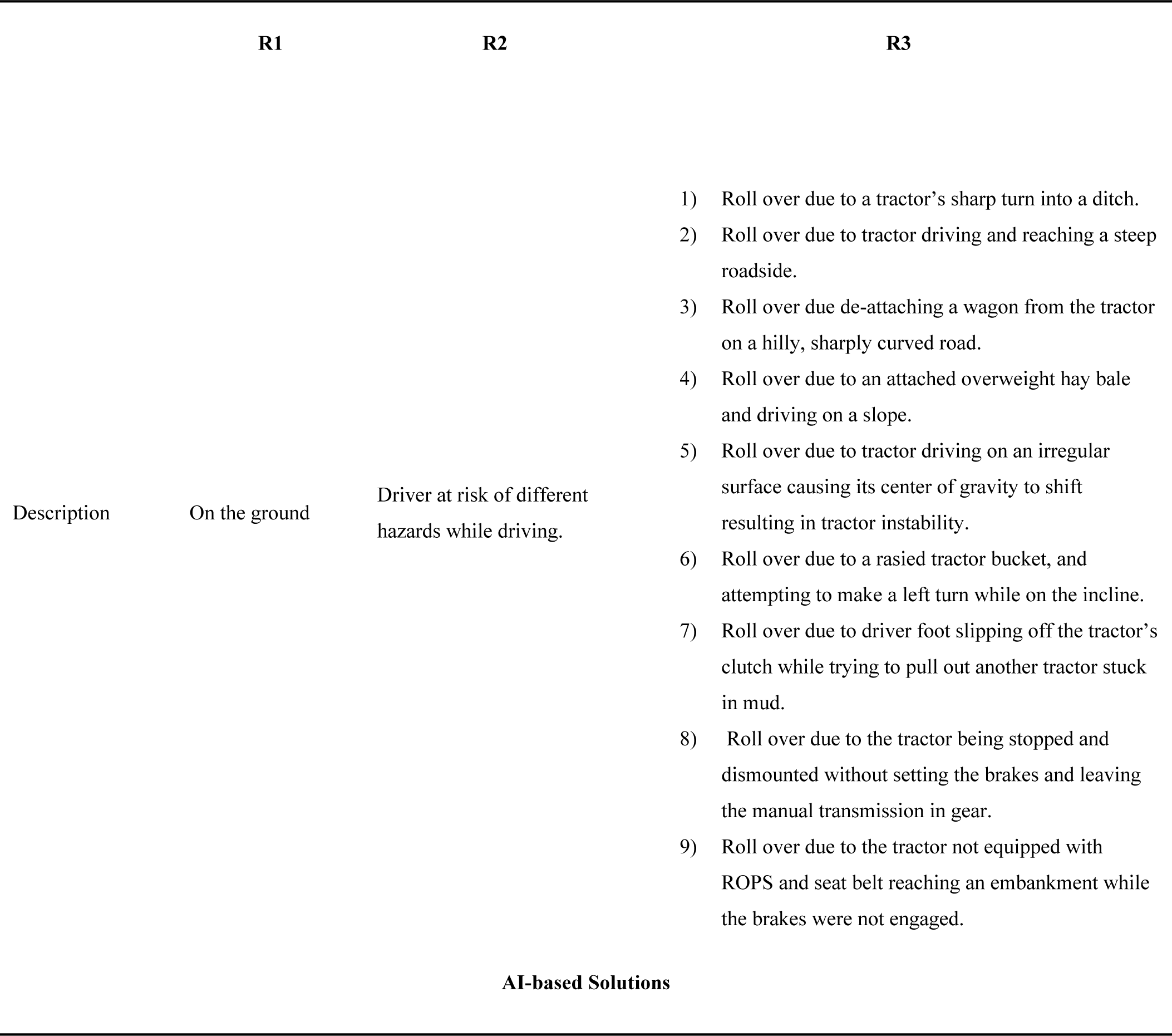
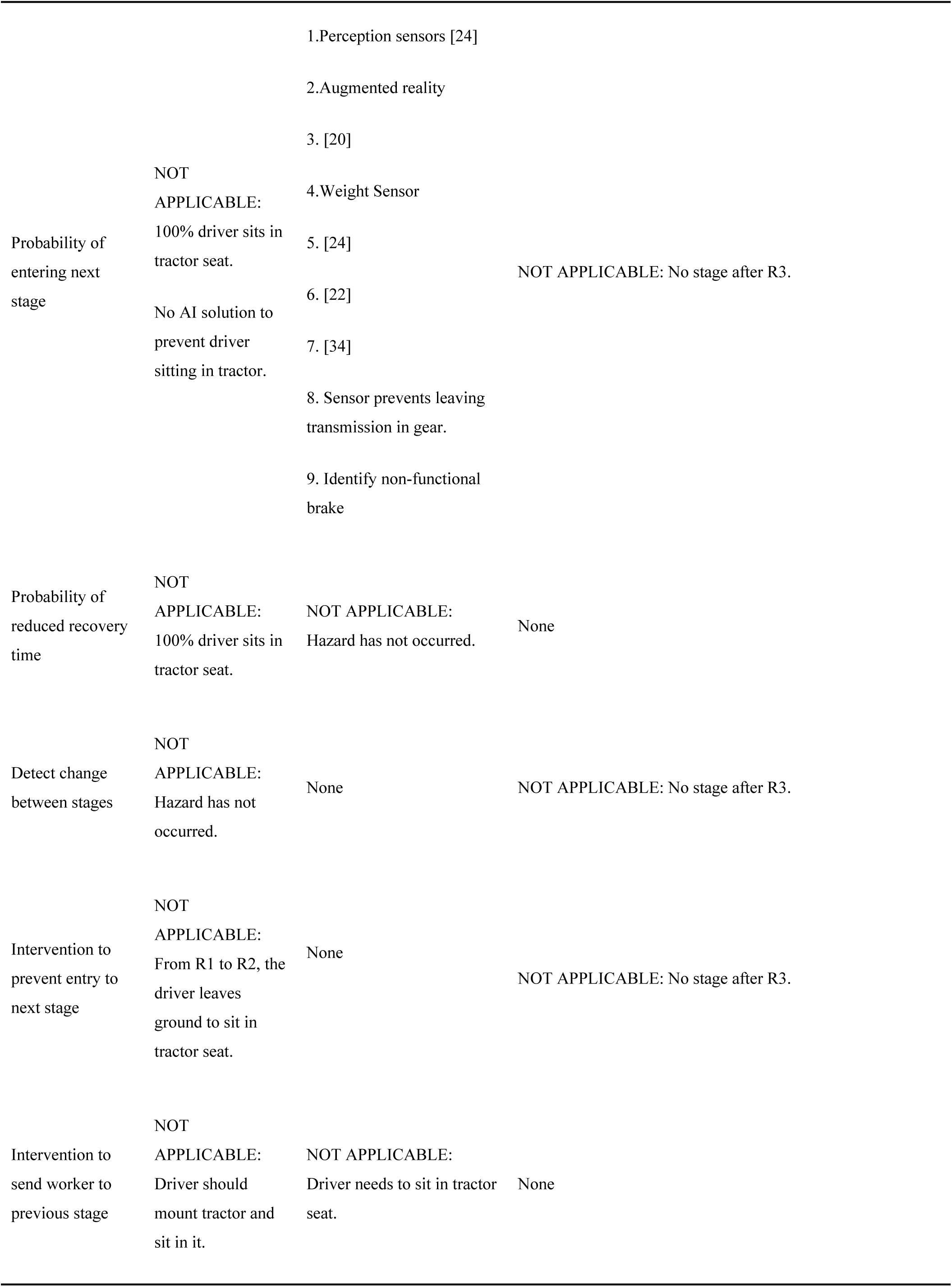
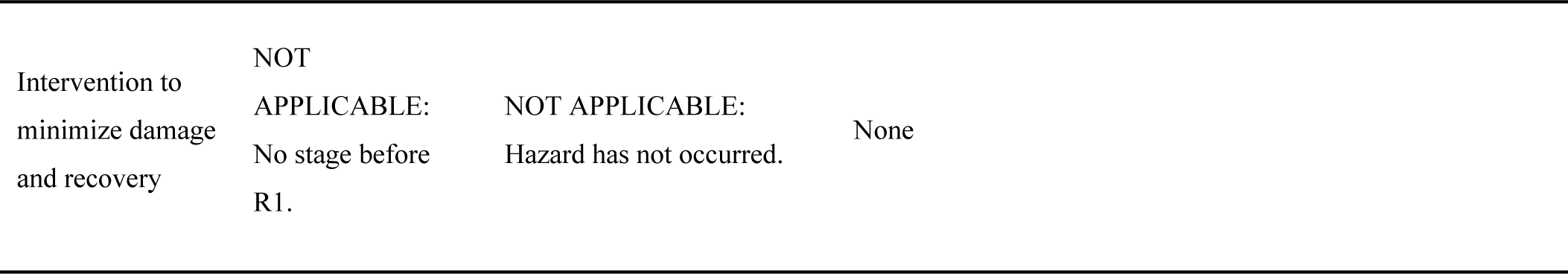
REDECA framework results summary for roll over cases.

**Table A.6.**
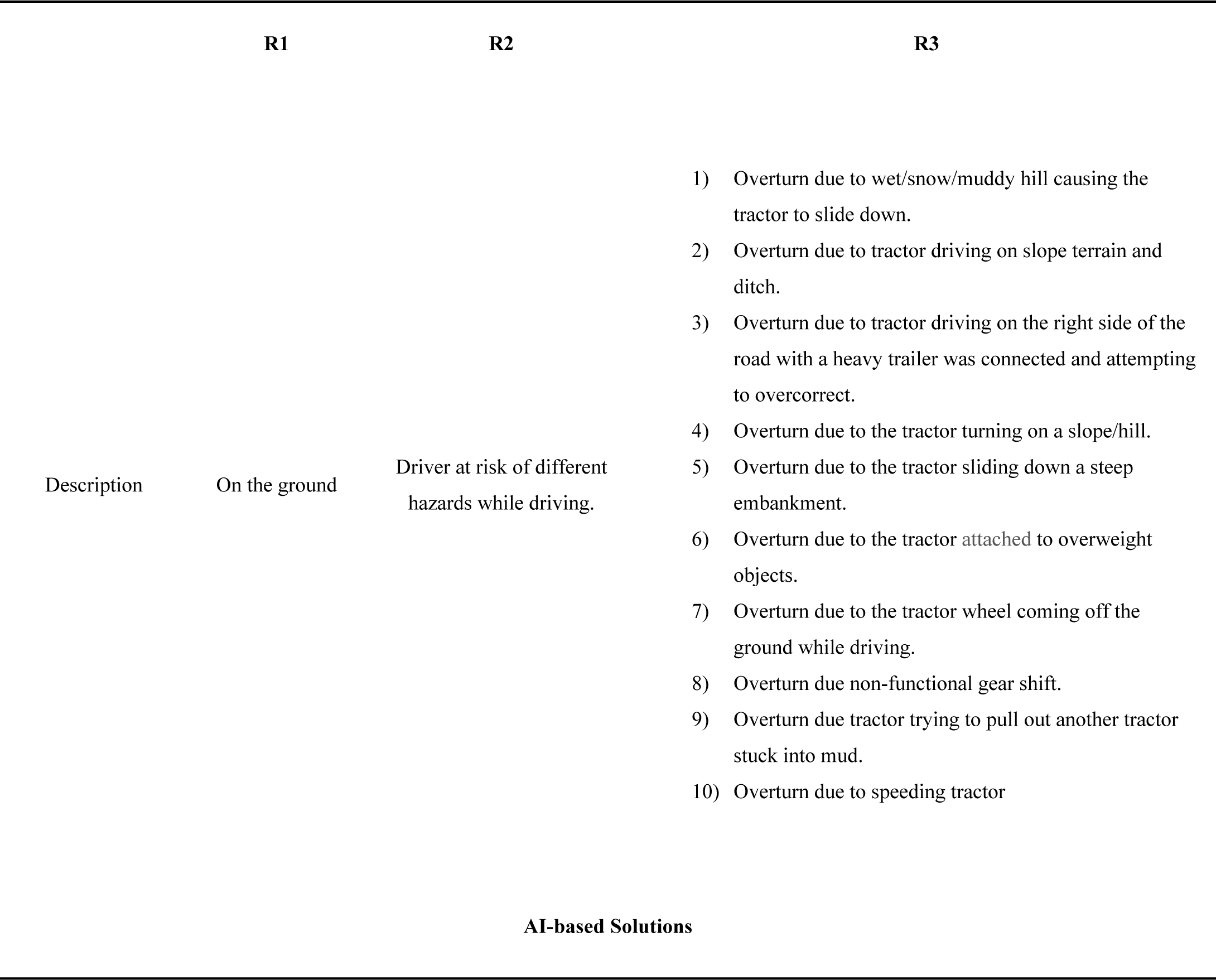
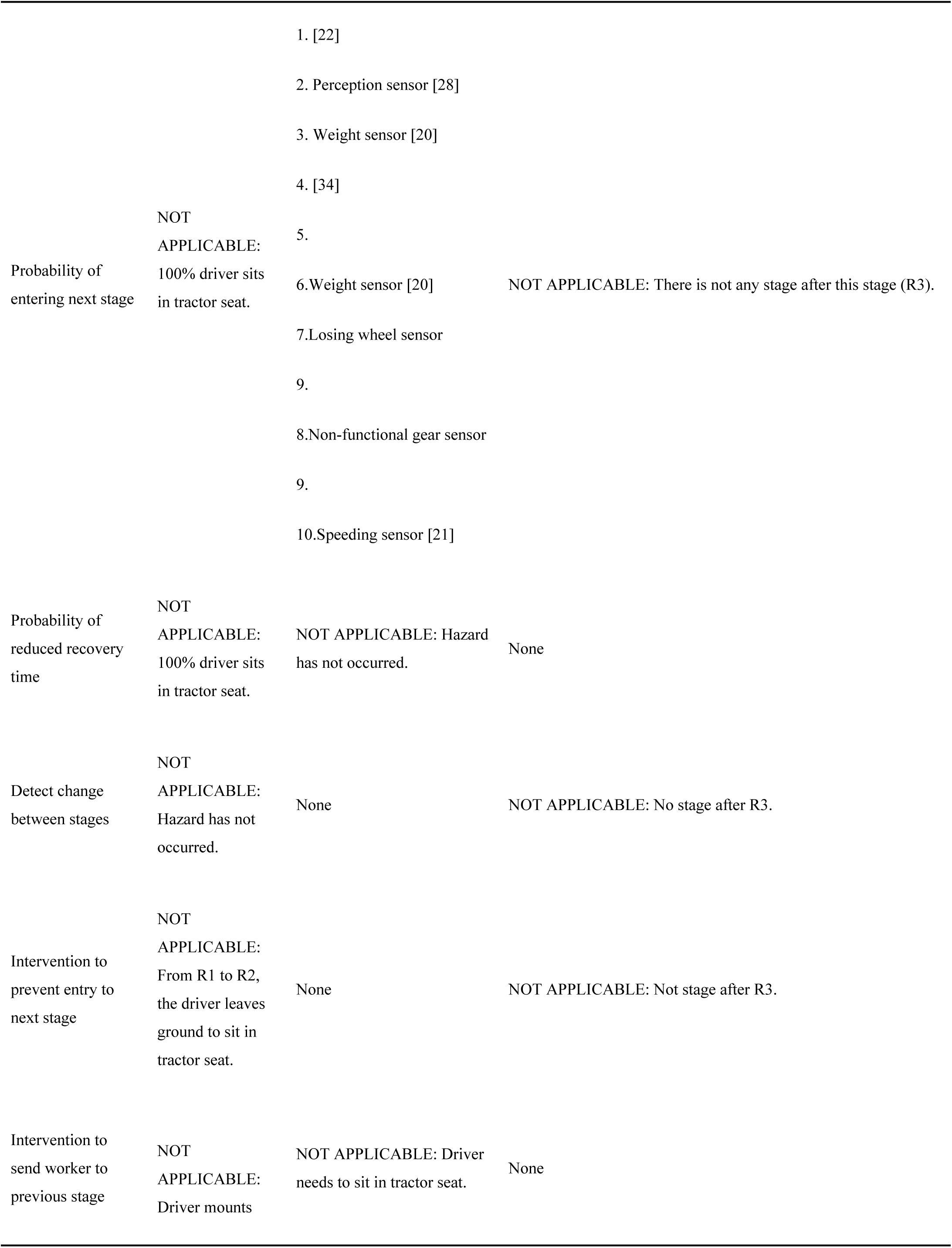
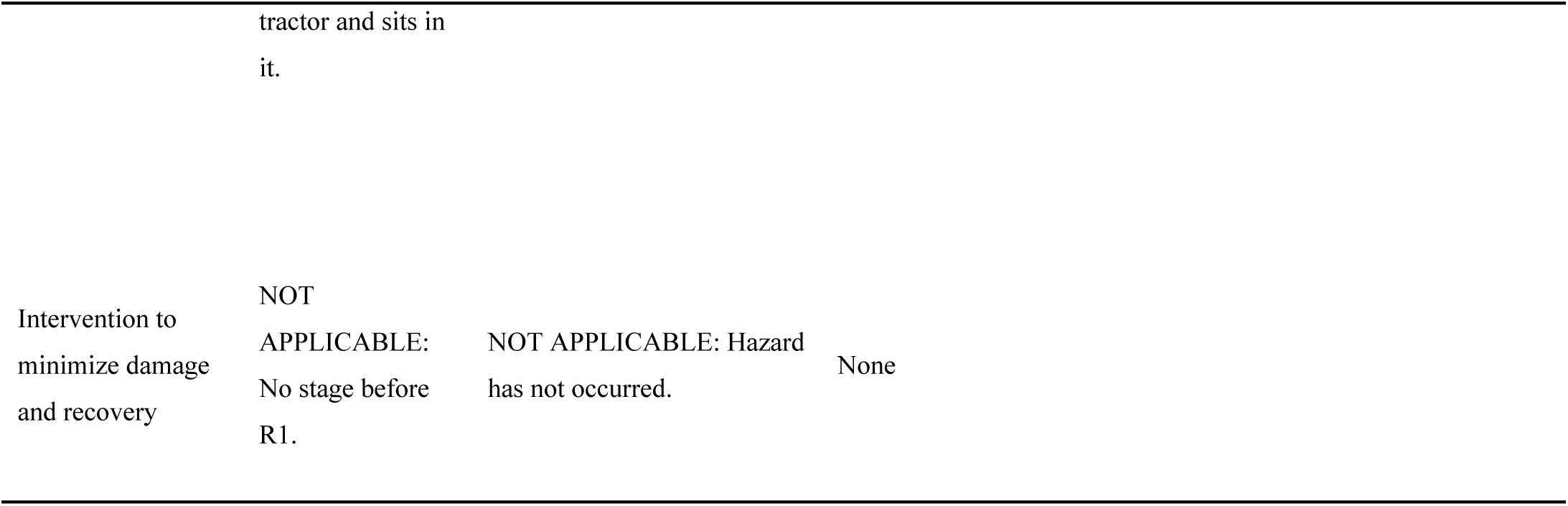
REDECA framework results summary for overturn cases.

